# Analysis of the SARS-Cov-2 epidemic in Lombardy (Italy) in its early phase. Are we going in the right direction?

**DOI:** 10.1101/2020.04.12.20062919

**Authors:** Angelo Riccio

## Abstract

**BACKGROUND:** We described the epidemiological features of the codiv-19 outbreak, and evaluated the impact of interventions measures on the epidemic in the Lombardy region, Italy.

**METHODS:** Laboratory-confirmed covid-19 cases reported through the beginning of April were extracted from the Italian Civil Protection database. Based on key events and interventions, we divided the epidemic into three periods: before February 21, from February 22 to early March, after early March. We compared epidemiological characteristics across periods and developed a modified susceptible-exposed-infectious-recovered model to study the epidemic and evaluate the impact of interventions. We explicitly took into account for unascertained cases (positive cases with no symptoms or mild symptoms that have not been accounted for in official statistics).

**RESULTS:** Currently, the number of positive active cases has increased to around 30,000 in the Lombardy region. Due to restriction measures, the effective reproduction number dropped from 3.33 (95% CI: 2.03–3.69) during the first period, to 2.36 (95% CI: 2.21–2.70) during the second period. In the third period, the effective reproduction number is estimated to have dropped to 1.49 (95% CI: 1.35–1.62). The model estimates a great proportion of unascertained cases, about 90% of infected people has not been accounted for in official statistics.

**CONCLUSIONS:** Considerable countermeasures have slowed down the covid-19 outbreak in the Lombardy region. However, notwithstanding the long-lasting lockdown period, the epidemic is still not under control. The effective reproduction number, according to the model used in this work, is still greater than 1.0. Estimation of unascertained cases has important implications on continuing surveillance and interventions.

## 1 Introduction

The novel coronavirus (SARS-CoV-2) emerged in Wuhan, China, in late 2019, and quickly spread rapidly to all Chinese provinces and other Asian countries (NHC, 2020). On March 11, 2020, the WHO (World Health Organization) declared the codiv-19 outbreak pandemic, after the disease caused by the new coronavirus infected more than 100,000 people and spread to more than 100 countries (WHO, 2020).

In the late night of February 20, 2020, the first case of novel coronavirus disease was confirmed in the Lombardy Region, northern Italy, around the city of Codogno. In the following week, the Codogno area, as well as several neighboring towns in southern Lombardy, experienced a very rapid increase in the number of detected cases, which rose to confirmed 172 positive samples by February 24 and 5,791 by March 10. From the early beginning of the outbreak, the Italian authorities have adopted a series of restrictive containment measures, including the creation of a ‘red zone’ around the city of Codogno and other small towns, progressively extended to several regions of northern Italy, and then, from March 10, also throughout Italy.

Efforts to contain the virus are ongoing; however, given the many uncertainties related to the transmissibility and virulence of this pathogen, the effectiveness of these efforts is unknown. Several recent studies have reported a nonnegligible proportion of asymptomatic cases and transmissibility of the asymptomatic or presymptomatic cases (Mizumoto et al., 2020; Li et al., 2020; Hu et al., 2020). Moreover, epidemiological data (dates of symptoms onset, clinical features, respiratory tract specimen results, hospitalization and contact tracing) collected by the Local Health Authority (ATS: Agenzia di Tutela della Salute) have allowed to establish that the epidemic in Italy began long before the clinical evidence of the first case ascertained in Codogno, when the virus had already spread to most of the municipalities of southern Lombardy (Cereda et al., 2020).

Here, to study the epidemic trend of this disease, we use a model inference to estimate the proportion of undocumented infections in the Lombardy region during the early phase of the epidemic, taking into account for intervention measures, ascertainment rate, transmission rate, and duration from illness onset to hospitalization. Model predictions have been compared against actual reported cases to evaluate the overall impact of interventions.

Using the data available in Italy, our objectives are to estimate:

- the number of people infected with covid-19 in the Lombardy region;
- the parameters of a SEIR-type model representing the early phase of the outbreak;
- the temporal modulation of the effective reproduction due to intervention measures.

## 2 Methods

### 2.1 Source of data

In Italy positive cases are first confirmed and validated in the laboratory and then distributed to the public through the Italian Civil Protection Department website (https://github.com/pcm-dpc/COVID-19). Positive cases are ascertained if the patient had a positive test of SARS-CoV-2 virus by the reverse transcription-polymerase chain reaction (RT-PCR) assay from the sequencing of nasal and pharyngeal swab specimens.

### 2.2 Classification of time periods

To better reflect the dynamics of the covid-19 epidemic and corresponding interventions, we classified the outbreak into three periods based on important dates that could affect the virus transmission:

1. the time before February 20;
2. the interval between February 20 and early March,
3. and the time after early March.

The first date corresponds the date of the first confirmed case in Italy, and this period was considered because no intervention was imposed before then. During the second period a series of progressively more stringent restriction measures have been adopted to contain the infection, precisely:

- February 24: closing of schools in Lombardy, Emilia Romagna and Vene-to regions;
- March 1: partial restriction of productive activities throughout Lombardy;
- March 5: schools closure throughout Italy;
- March 8: lockdown of mobility in Lombardy.

After March 10, a severe lockdown of almost all activities has been extended to the whole of Italy.

### 2.3 Statistical analysis

Due to the severe shortage of medical resources early on in this epidemic, many suspected cases were unable to receive timely treatment and were self-quarantined at home. It is also believed that many deaths related to the covid-19 epidemic have not been correctly accounted for in official statistics, so that the real extent of the epidemic has been probably underestimated in Lombardy.

To infer SARS-CoV-2 transmission dynamics during the early stage of the outbreak, we simulated observations starting from February 24, 2020 (i.e., when data from the Italian Civil Protection were made available), extending the classic SEIR model to account for quarantine measures and unascertained cases. We divided the population into six compartments including susceptible individuals, exposed cases, ascertained cases, unascertained cases, hospitalized cases, and recovered individuals. Here, unascertained cases included asymptomatic cases and those with mild symptoms that could recover without seeking medical care and thus were not reported to the authorities. The model dynamic is illustrated in Figure 1.

**Figure 1:**
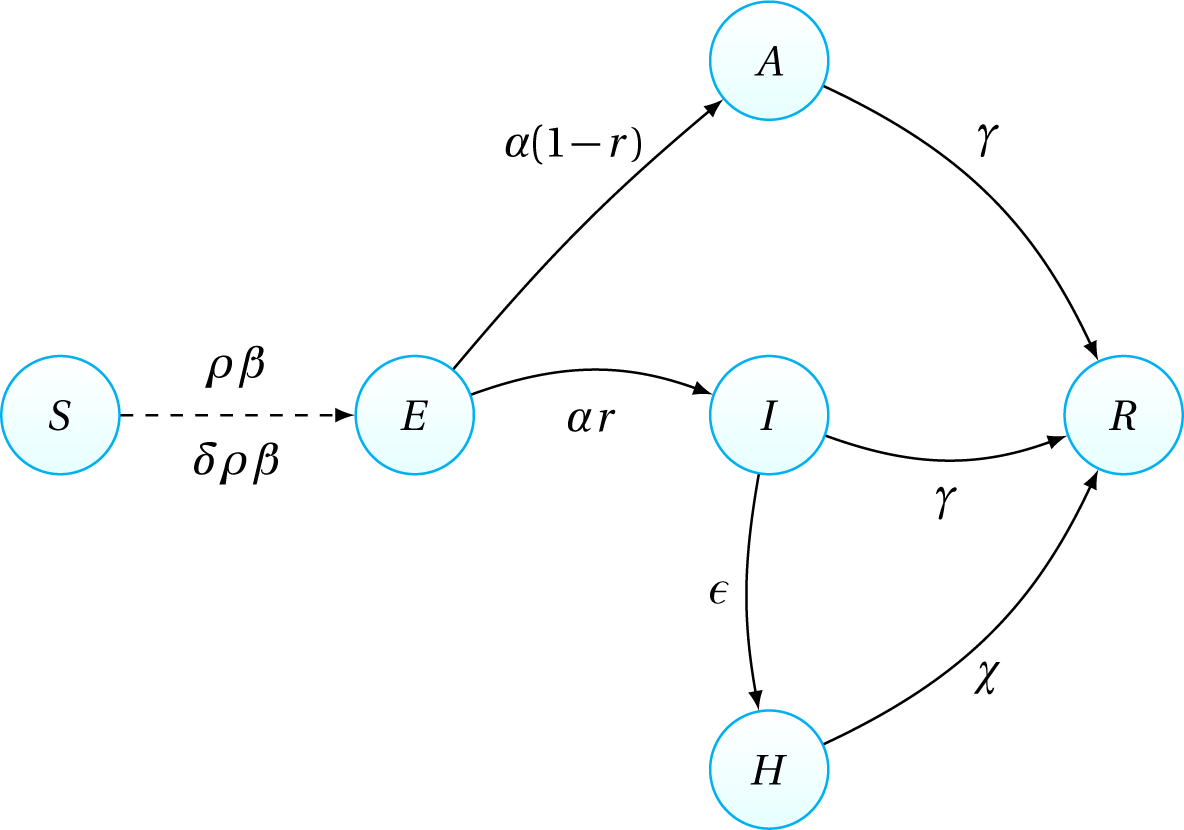
Illustration of the extended SEIR model. The symbols for compartments are: *S* (susceptible), *E* (exposed), *I* (reported infectious), *A* (unreported infectious), *H* (hospitalized), and *R* (recovered). The rate associated with each path is indicated by the corresponding basic parameter.

The key parameters in the model are:

1. *ρ* (effect of intervention measures),
2. *r* (ascertainment rate), and
3. *β* (transmission rate).

For unreported infectious individuals, the transmission rate may be different from that of infectious; this is taken into account by the parameter *δ*.

Considering the impacts of major interventions, we allowed the reproduction number to vary over time, rather than simply fix this value, to capture possible variation in transmission as a result of control measures and behaviour change. We assumed that the ascertainment rate and transmission rate were different for the three time periods. The effective reproduction number, *R*_*t*_, defined as the expected number of secondary cases infected by a primary case, was computed for each period. Initial states and parameter settings of the SEIR model are described in detail in the Appendix A.

## 3 Results

The epidemic curve is illustrated in Figure 2. During the first phase, the infection curve grew rapidly and, despite containment measures, no significant slowdown was observed. For this reason, on 8 March it was decided the lock-down of all activities. Only after several days, the infection curve began to show a slowdown. Currently, on 10 April, the number of people infected in the Lom-bardy region is about 30,000.

**Figure 2:**
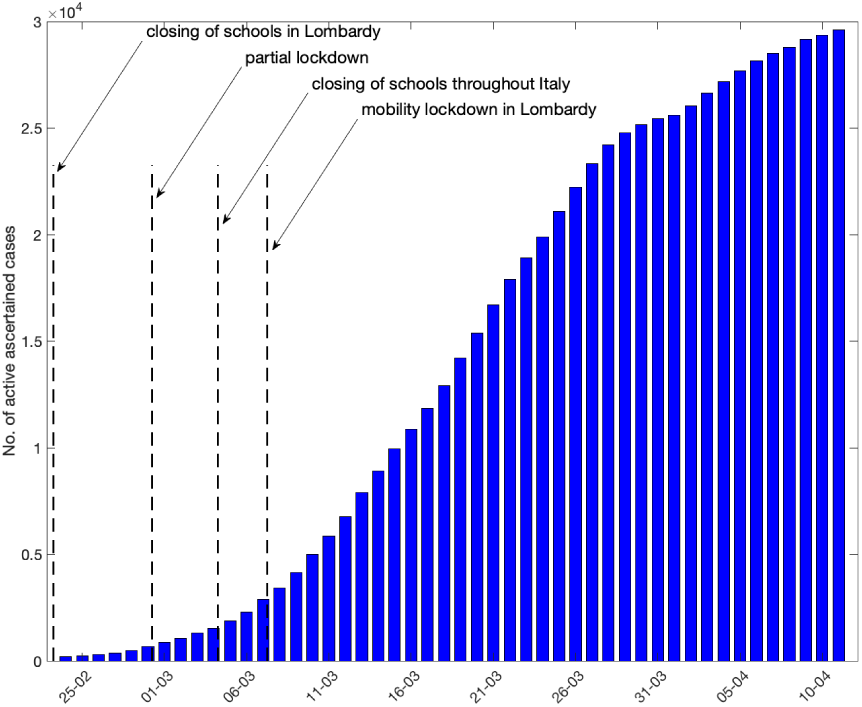
The daily number of ascertained cases. The vertical black dashed lines are drawn at the time when the different control measures where taken.

Our SEIR model fit the observed data reasonably well (Figure 3). To adapt to the observed data, the model estimates that it is possible to date the beginning of the epidemic to the first decade of January, precisely to 10 January (95% CI: 5–14 January). In addition, the effectiveness of lockdown, namely the effects of the progressive restriction measures, started to show their effects on 17 March (95% CI: 14–19 March).

**Figure 3:**
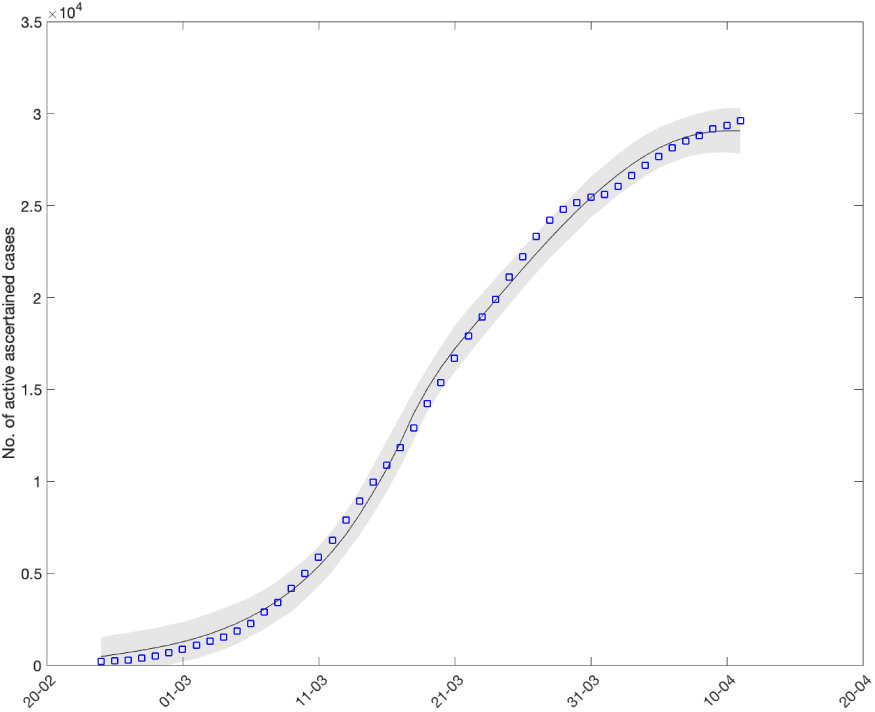
Fit of the SEIR model to the number of active ascertained cases in Lombardy. The grey shaded area represents the 95% CIs.

Strikingly, we estimated that the overall ascertainment rate was 0.13, and similar across the last two periods. Precisely, during the second period the median value of the ascertainment rate, *r*_1_, is 0.13 (95% CI: 0.10–0.17) and during the second period the ascertainment rate, *r*_2_, is 0.13 (95% CI: 0.11–0.17), that is, about 90% of infected people has not been included in official statistics, according to our SEIR model. During the first days of April 2020, we estimate about 250,000 unascertained cases in Lombardy, i.e. about 2.5% of the whole population in this region, see Figure 4. Only 30,000 cases have been accounted for in official statistics.

**Figure 4:**
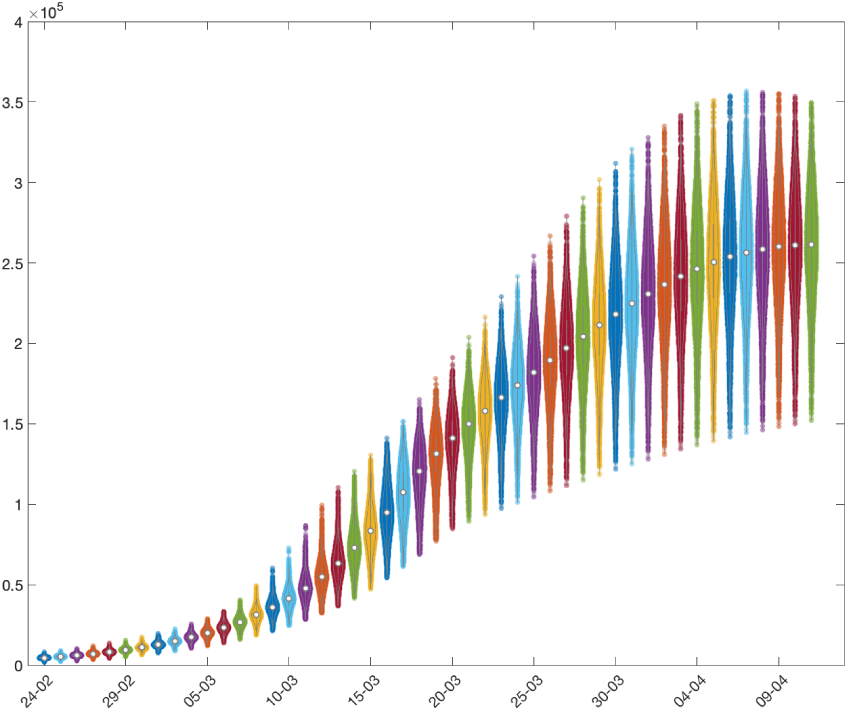
Number of unascertained cases in Lombardy.

The estimated effective reproduction number, *R*_*t*_, is shown in Figure 5. During the first period, i.e. before February 24, when no control measure where taken, the reproduction number is estimated to be 3.33 (95% CI: 2.03–3.69), and slowed down to 2.36 (95% CI: 2.21–2.70) during the second period. Un-fortunately, it should be noted that the curve of active cases is slowing down slowly. This feature translates into an *R*_*t*_ value which is still relatively high. In the third period it is estimated that *R*_*t*_ is greater than 1, precisely 1.49 (IC 95%: 1.35-1.62), despite the length of the lockdown that, on April 10, lasts for more than a month.

**Figure 5:**
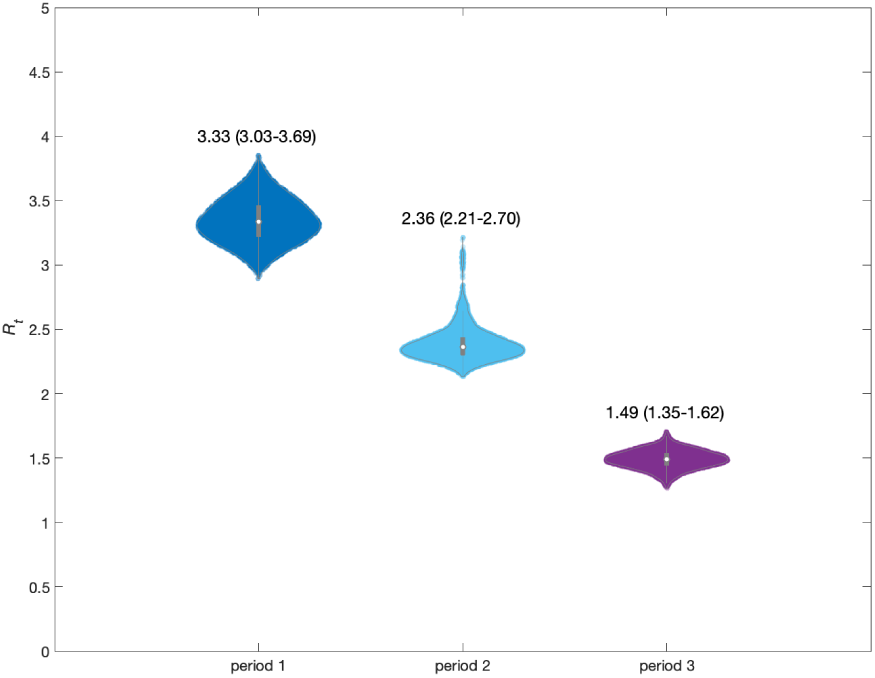
Estimated *R*_*t*_ for the three periods. For each time period, the median and the 95% CI are reported above the violin plot.

As more data become available on the timing of control measures and sub-sequent dynamics, we agree that it will be crucial to evaluate the effectiveness of measures to provide a robust evidence base for future policy-making.

## Data Availability

All data used in this work are public available

https://github.com/pcm-dpc/COVID-19

## A Model description

We extended the classic SEIR model to account for unascertained cases and quarantine measures (Figure 1). We divided the population into *S* (susceptible), *E* (exposed), *I* (ascertained infectious), *A* (unascertained infectious), *H* (hospitalized) and *R* (recovered) individuals.

Starting with a total population size of 10.06 million (equivalent to that of the Lombardy region) and 1 exposed individual, the model update the number of each compartment, according to the following set of ordinary differential equations:

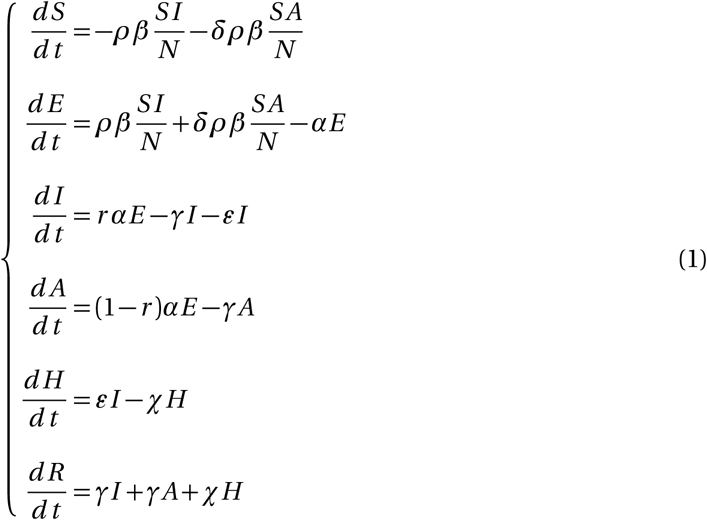

The initial state of the model is reported in Table 1.

**Table 1:** Initial state for the *S* and *E* compartments of the SEIR model. All other initial values are set to 0

| Parameter | meaning | before |
| --- | --- | --- |
| $S_0$ | transmission rate | 10.06M |
| $E_0$ | Number of exposed individuals | 1 |

*β* is the basic reproduction number, i.e. the transmission rate without any intervention measure, defined as the number of individuals that a positive case can infect per day; *ρ* weights the effect of intervention measures; *δ* is the ratio of the transmission rate of unascertained over ascertained cases; *r* the ascertainment rate; 1*/α* and 1*/γ* are the latent and infectious time scale; 1*/ε* is the time scale from illness onset to hospitalization and 1*/χ* the hospitalization time scale. Our model was similar to that in another study, which focused on case counts in Wuhan (Wang et al., 2020).

The SEIR model is started at *t* = *t*_0_, which should approach the date of introduction of the virus. Here we assume that *t*_0_ is unknown and is a parameter estimated by the model. Because of the large number of intervention measures taken, it is not clear at what time they have become effective. In the model, *t*_2_, the starting time of the third phase, was considered unknown, and estimated by the data themselves.

For this model the effective reproduction number, *R*_*t*_, can be calculated as

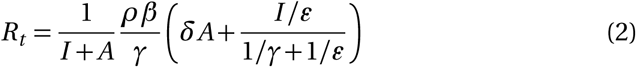

*R*_*t*_ depends on time in this model, because *I* and *A* depend on time and because the effectiveness of intervention measures, i.e. the *ρ*s, depends on time. We took the mean across time within a given period as the estimate of *R*_*t*_.

We informed model parameters with estimates from other countries, where available (Table S1, Hellewell et al. (2020)), and estimated the values of unknown parameters by fitting the model to data on local reported positive cases in the Lombardy region (see Table 2).

**Table 2:** Parameters and model values of the SEIR model for the three periods

|  |  | period |  |  |
| --- | --- | --- | --- | --- |
| variable | meaning | 1 | 2 | 3 |
| $t_0$ | starting time of infection | | $t_0$ | |
| $t$ | starting time of each period (days from January 1) | 0 | 54 | $t_2$ |
| $\beta_0$ | basic reproduction rate | $\beta_0$ | $\beta_0$ | $\beta_0$ |
| $\rho$ | intervention effect | 1 | $\rho_1$ | $\rho_2$ |
| $r$ | ascertainment rate | 0 | $r_1$ | $r_2$ |
| $\delta$ | ratio of unascertained/ascertained transmission rate | 1 | 1 | 1 |
| $\alpha$ | latency rate (days<sup>-1</sup>) | 0.19 | 0.19 | 0.19 |
| $\gamma$ | infection rate (days<sup>-1</sup>) | 0.43 | 0.43 | 0.43 |
| $\varepsilon$ | rate from illness onset to hospitalization (days<sup>-1</sup>) | 0.14 | 0.14 | 0.14 |
| $\chi$ | rate of hospitalization (days<sup>-1</sup>) | 0.033 | 0.033 | 0.033 |

To estimate the unknown parameters (*t*_0_, *β*_0_, *ρ*_1_, *ρ*_2_, *r*_1_ *r*_2_ and *t*_2_), we assumed that the number of ascertained cases estimated by the model with illness onset on day *t*, denoted as *y*_*t*_, follows a Poisson distribution with rate parameter *λ*_*t*_, *y*_*t*_ *∼* Poisson(*λ*_*t*_), where *λ*_*t*_ is the number of notified positive cases on day *t*. Thus the likelihood function was

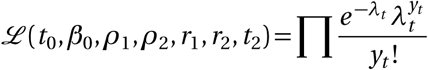

In order to compute the *a-posteriori* estimator (i.e., the parameters that maximize the posterior function), we used rather vague priors and imposed the following feasibility intervals to sample from:

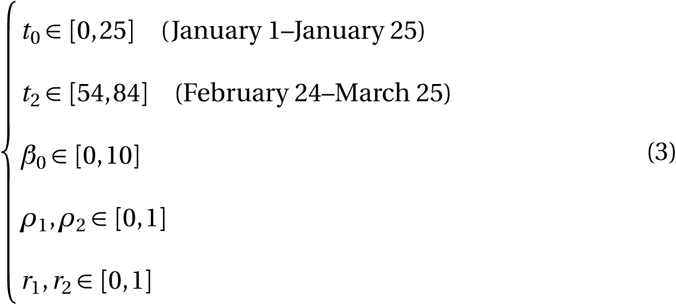

The numerical computation of the posterior distribution is performed with a Metropolis-Hastings (MCMC) algorithm from 100,000 iterations, using the DRAM algorithm (Delayed Rejection Adaptive Metropolis, Haario et al. (2006)). For prediction, we obtained CIs (Credible Intervals) by stochastic simulations under the SEIR model with the sampled parameter values from MCMC. The *a-priori* and *a-posteriori* probability density functions are shown in Figure 6.

**Figure 6:**
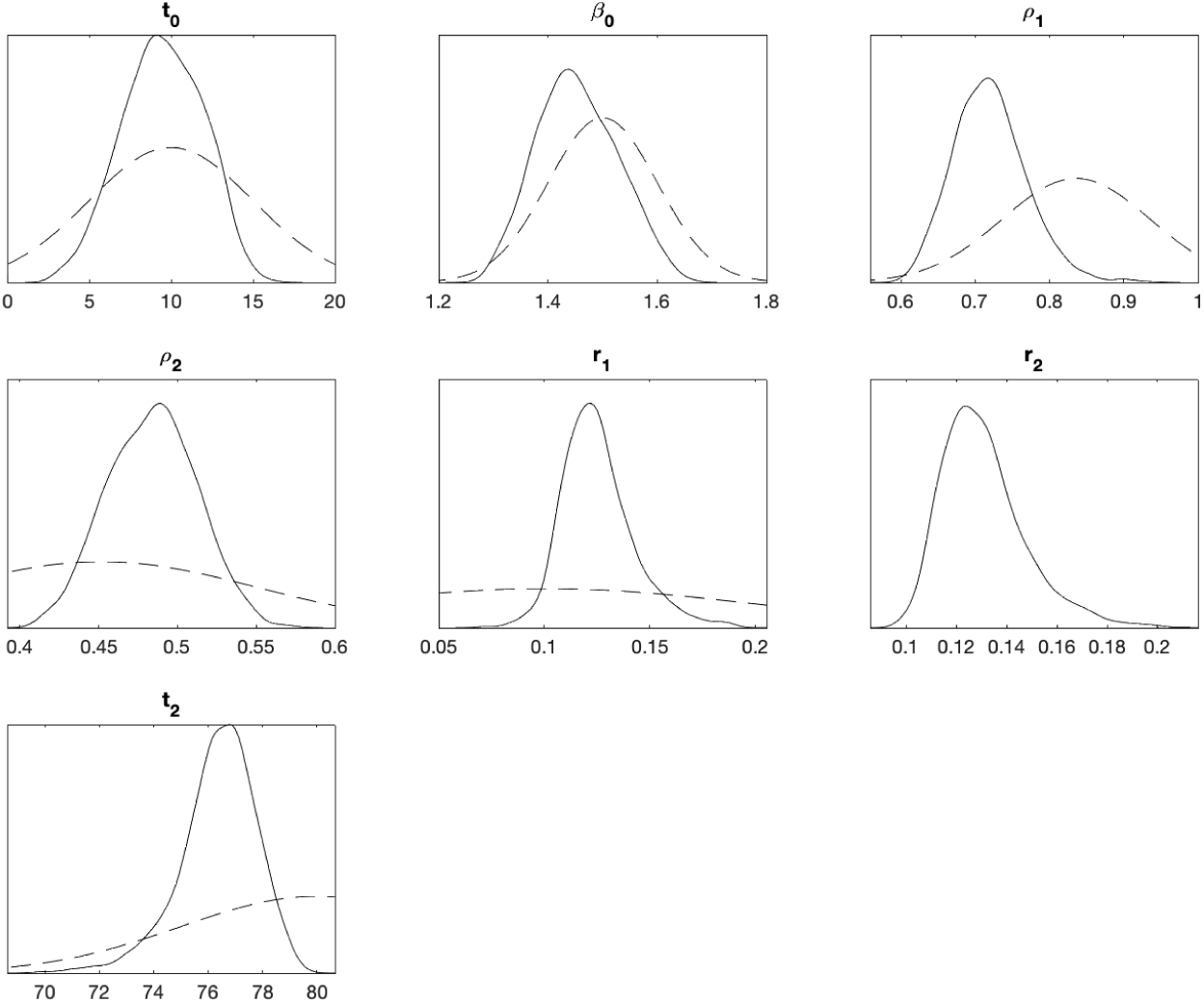
*a-priori* (dashed lines) and *a-posteriori* (solid line) probability density functions for all parameters of the SEIR model.

## Notes

### Competing Interest Statement

The authors have declared no competing interest.

### Funding Statement

We declare that no funding has been received to support the analysis presented in this manuscript.

